# A blind trial of Aptamarker prediction of brain amyloid based on plasma analysis

**DOI:** 10.1101/2023.11.15.23298582

**Authors:** Soizic Lecocq, Filipa Bastos, Mariana Silva, Cathal Meehan, Rita Castro, João Cunha, Ana Cristina Silva, Luis Ruano, Gregory Penner

## Abstract

We previously described the development of a predictive blood test for brain amyloid based on the combined use of eight aptamers (Aptamarkers) and the clinical variable, patient age with a sensitivity of 88%, specificity of 76% and an overall accuracy of 81%. This test results from an agnostic application of a defined library of 4.29 billion aptamer sequences to plasma samples derived from individuals with varying amounts of brain amyloid deposition. In this report, the same eight Aptamarkers and corresponding predictive algorithm were applied to plasma samples from 36 patients diagnosed with memory impairment in a memory clinic in Portugal. The patients were all subjected to cerebrospinal fluid (CSF) analysis with Aβ40, Aβ4 peptides, p181-tau and total tau characterization. The plasma analysis with the Aptamarkers resulted in a sensitivity of 73%, specificity of 71% and overall accuracy of 72% when considering brain amyloid alone. The predictive power when considering the diagnosis of Alzheimer’s disease based on a complete CSF analysis was a sensitivity of 68%, a specificity of 76% and an overall accuracy of 72%. These results in this diagnostic accuracy study are very similar to results obtained with model building on AIBL cohort. As such, this study demonstrates the potential efficacy of this simple qPCR assay as a basis for inclusion for subsequent CSF confirmation of brain amyloid deposition in a clinical setting.

## Background

Clinicians at memory clinics are presented with patients with memory complaints of uncertain pathophysiological cause. The assignment of an individual pathophysiological basis for these symptoms is critical for the appropriate clinical care and for the planning of necessary support by care-givers. To determine whether a patient is affected by Alzheimer’s disease (AD), clinicians can choose to extract cerebrospinal fluid (CSF) with a lumbar puncture and analyze the fluid for the ratio between Abeta-40 and Abeta-42 peptides^1^ and phospho-tau versus total tau protein^2^. It would be beneficial if a first line blood-based test existed to identify individuals at risk for Alzheimer’s disease^3,4^. In the event where first line blood tests indicate high brain amyloid deposition, patients would be subjected to a lumbar puncture and the ratios of CSF biomarkers would be ascertained to facilitate an AD diagnosis. Conversely, first line blood tests which predict low brain amyloid would exclude some patients from subsequent CSF analysis. The invention of a first line blood-based test would ultimately reduce the number of invasive CSF tests performed on individuals who are unaffected by AD. The elimination of unnecessary CSF tests would alleviate the burden on hospital staff and decrease total healthcare expenditure.

Recently there has been an emergence of biomarkers identified in plasma including Aβ peptides, phosphor-tau and neurofilament light chain protein (NFL). First line analyses for these biomarkers conducted on afflicted individuals are primarily becoming available through Simoa platforms. Unfortunately, the sophistication of the equipment required for these tests necessitates central laboratory delivery, which increases expenses and complicates blood deliveries transported from clinics to testing laboratories. An urgent need currently exists for a simple plasma test to assist clinicians with pathology assessments. A routine plasma test would enable in house testing, with existing equipment and expertise.

In response, NeoNeuro has developed what they refer to as the Aptamarker platform^5^. The aptamarker platform is derived from a common library of the same 4.29E9 aptamer sequences is applied to blood samples from individuals that differ in phenotype. This approach has been used in conjunction with blood samples and data from the AIBL cohort on ten individuals with high brain amyloid, and ten individuals with low brain amyloid. Differences in the frequencies of the aptamers between the two phenotypic pools was evaluated, and a subset of eight predictive aptamers was identified. These aptamers are now referred to as Aptamarkers. Primers were designed for each Aptamarker to enable qPCR quantification of each sequence independently. A predictive algorithm was developed based on the binding of these Aptamarkers to their targets in 391 blood samples along with the clinical variable age. The trained model resulted in a sensitivity of 88%, specificity of 76% and an overall accuracy of 81%.

This test and an accompanying predictive algorithm were applied to plasma from 36 individuals all diagnosed with mild cognitive impairment in a diagnostic accuracy study with a complete CSF analysis as the golden standard.

## Methods

### Clinical setting

The study was performed on a sample of patients recruited from Memory Outpatient Clinic of Centro Hospitalar de Entre Douro e Vouga (CHEDV), CHEDV is a 400-bed hospital located in Northern Portugal, with a reference population of 360.000 people within the Portuguese NHS. Two groups were selected for this study. The first group contained patients with an Alzheimer’s disease diagnosis, confirmed by CSF biomarkers. The second group of patients was a control group of patients with suspected/diagnosed dementia with diagnosis of Alzheimer’s disease excluded by CSF biomarkers.

### Patient inclusion criteria

All participants were > 43 years old and had a clinical dementia rating of 1.0 or lower (Except for one patient that exhibited a clinical dementia ratio of 3 and was diagnosed with Parkinson’s disease). Patients who were subjected to CSF analysis in the second semester of 2023 and complied with inclusion criteria were consecutively invited to participate in this study. All the participants or their legal representatives provided written consent for the use of their medical information in this study.

Inclusion criteria for the Alzheimer’s disease group

1. Progressive cognitive deterioration with a clinical diagnosis of AD, according to the guidelines of the National Institute on Aging-Alzheimer’s Association (McKhann GM et al, 2011)
2. Positive neurochemical molecular diagnosis,
3. Magnetic resonance imaging (MRI) or Computed Axial Tomography (CT) of the brain not suggestive of an alternative diagnosis.

Inclusion criteria for the control group

1. Suspected cognitive impairment and/or confirmed diagnosis of dementia other than Alzheimer’s disease
2. With CSF biomarkers clearly negative, both beta-amyloid above normative values and phospho-Tau-181 below normative values
3. No typical findings of AD on brain MRI/CT scans (mesial temporal atrophy or parietal atrophy).

A total of 42 consecutive patients that subjected to CSF analysis for Alzheimer’s disease between July and December 2023 were invited to the study, with 36 accepting the invitation. Reasons for refusal were the difficulty to visit the hospital for the sample collection in the scheduled days.

### Samples

The samples were shared with NeoNeuro in a blinded manner with no reference to clinical variables other than sex and age. Heparin prepared plasma was shipped from CHEDV to

NeoNeuro for analysis. It should be noted that the ice had melted by the time the samples were received by NeoNeuro. Plasma was subjected to at least three freeze thaw cycles prior to analysis.

### DNA library and primers

The eight Aptamarkers used on the AIBL cohort^8^ were used in the same manner for this analysis. The sequences of these eight Aptamarkers are provided in Table 1. Specific reverse primers were designed for each Aptamarker. A common forward primer was designed for use across all Aptamarkers: 5’-GTT CCC AGA TAC AGA C-3’. Primers were tested to ensure that each set specifically amplified the targeted Aptamarker sequences using a melt curve. The melt curve protocol followed with 10 seconds at 95°C and then 5 seconds each at 0.5°C increments between 55°C and 95°C. Data collection was enabled at each increment of the melt curve. Table 1 details the specific sequences of the Aptamarkers used in the study, alongside their corresponding reverse primer sequences and concentrations.

**Table 1:**
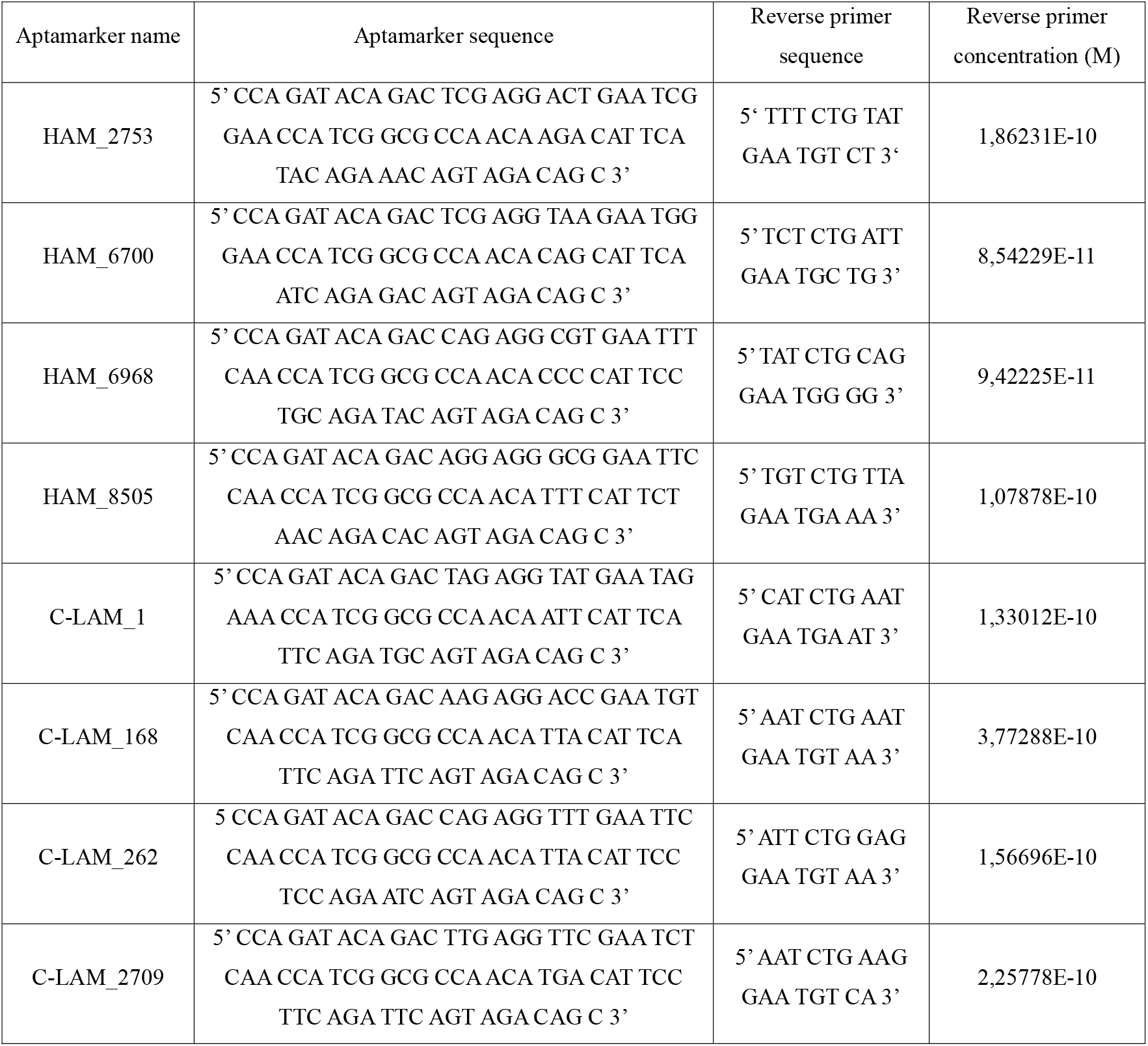
Sequences and Concentrations of Aptamarkers and Reverse Primers Used in the AIBL Cohort Analysis.

### Antisense design

A universal antisense oligonucleotide was designed with the capacity to hybridize to each of the eight Aptamarkers. The antisense oligonucleotide was ordered from Integrated DNA Technologies (IDT) with a thiol modifier on the 5’-terminus in order to conjugate the antisense to a solid gold support during selection. The sequence of the antisense oligonucleotide was /5ThioMC6-D/AAAACAAAGTCTACTTGTTGGTTCTGTAT 3’..*

### Gold nanoparticle conjugation

The thiol modifier on the universal antisense oligonucleotide was reduced with tris (2carboxyethyl) phosphine (TCEP) following IDT-DNA specifications. The reduced antisense was conjugated to gold nanoparticles (GNPs) (40nm diameter, Cytodiagnostics) following supplier specifications. GNPs were suspended in 200 µL 0.1X of phosphate buffer saline (PBS) (0,8 mM Na_2_PO_4_, 0,14 mM KH_2_PO_4_, 13,6 mM NaCl, 0,27 mM KCl, pH 7.4). The amount of antisense was evaluated before and after conjugation using a nanodrop. A final concentration of 240 nM antisense was used in all experiments in a volume of 50 μL.

### Selection

The eight Aptamarkers were pooled at the concentrations indicated in Table 1. The pooled Aptamarkers were incubated with a 10 μL volume of plasma obtained from the CHEDV in a 20 mM Tris buffer containing 120 mM NaCl, 5mM MgCl_2_, 5 mM KCl were incubated with 10 μL of plasma extracted from patients with mild cognitive impairment received from CHEDV. The final concentration of buffer in this 50 uL solution was 20 mM Tris, 120 mM NaCl, 5 mM MgCl_2_, 5 mM KCl. Samples were incubated with functionalized GNPs for 15 minutes and were centrifuged for 20 minutes at a speed of 3,500 RCF. Following centrifugation, 30 µL of the supernatant was collected and diluted with 80 µL of sterile water. The pellet was discarded.

### qPCR analysis

These aptamers were analyzed by real-time PCR (qPCR) using the primers referred to in Table 1. 5 uL of the diluted supernatant was used as template for each Aptamarker. Quantification of the qPCR amplification was achieved through the inclusion of a SYBR green dye according to the manufacturer’s instructions (Bio-Rad). Each 20 μL reaction mix contained 5 μL of extracted DNA, 250 nM PCR primers and 1 X SYBR green master mix. The reaction was initially denatured at 95°C for 5 minutes followed by 35 cycles of amplification (95°C for 10 seconds, 40.6°C for 15 seconds and 72°C for 30 seconds).

### Data analysis

The raw data from the qPCR assays was analyzed in the same manner as described for the AIBL cohort study^5^ to generate relative template proportion values (RTp). These values were preprocessed using the same scaling as on the AIBL training data. High brain amyloid deposition in patients was classified where the Abeta_42/Abeta_40 ratio was below a threshold of 0.05.

Amyloid status for the CHEDV dataset was predicted utilizing the previously identified model based on an ExtraTrees algorithms described for the analysis on the AIBL dataset^6^.

## Results

The CHEDV analysis can be separated into two relevant categories for this study, Abeta_42/Abeta_40 peptide ratios as an indication of brain amyloid deposition, and diagnosis of Alzheimer’s disease based on a combination of this ratio and P-tau data. A summary of individual patient clinical and demographic characteristics is provided in supplementary data. All patients were classified as Portuguese in ethnicity. Twenty two of the 36 individuals were female at birth. The average age was 67, with the youngest being 43 and the oldest 82. 19 of the patients were diagnosed with AD, 9 with depression, and the others with a variety of pathophysiological causes. Those collecting the blood samples and carrying out the diagnostic test were blinded to the brain amyloid deposition classification of the patients. The diagnostic test prediction was carried out using qPCR values from the 8 Aptamarkers selected in blood samples and the age of the patient.

We compared the Aptamarker predictions to the Abeta_42/Abeta_40 ratio results with a definition that a ratio less than 0.05 indicated high brain amyloid accumulation. The data for this comparison is provided in Table 2 and a confusion matrix summarising the data is provided in Table 3. The table compares the predictive values obtained from Aptamarker analysis with the Abeta_42/Abeta_40 ratio results. A threshold ratio of less than 0.05 was used to indicate high brain amyloid deposition.

**Table 2:**
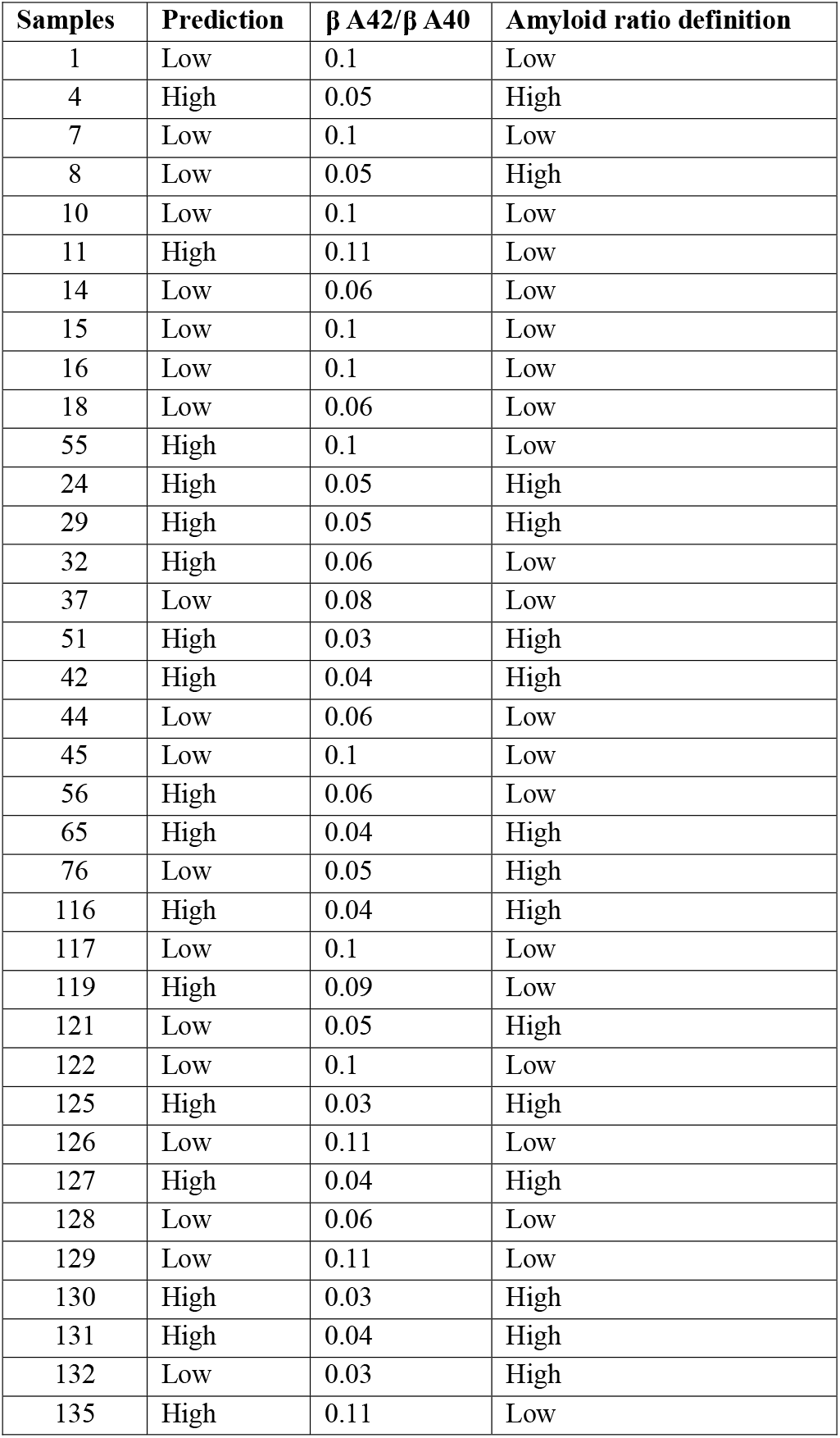
Comparison of aptamarker predictions with Abeta_42/Abeta_40 ratio for indicating brain amyloid deposition.

| Samples | Prediction | $\beta$ A42/$\beta$ A40 | Amyloid ratio definition |
| --- | --- | --- | --- |
| 1 | Low | 0.1 | Low |
| 4 | High | 0.05 | High |
| 7 | Low | 0.1 | Low |
| 8 | Low | 0.05 | High |
| 10 | Low | 0.1 | Low |
| 11 | High | 0.11 | Low |
| 14 | Low | 0.06 | Low |
| 15 | Low | 0.1 | Low |
| 16 | Low | 0.1 | Low |
| 18 | Low | 0.06 | Low |
| 55 | High | 0.1 | Low |
| 24 | High | 0.05 | High |
| 29 | High | 0.05 | High |
| 32 | High | 0.06 | Low |
| 37 | Low | 0.08 | Low |
| 51 | High | 0.03 | High |
| 42 | High | 0.04 | High |
| 44 | Low | 0.06 | Low |
| 45 | Low | 0.1 | Low |
| 56 | High | 0.06 | Low |
| 65 | High | 0.04 | High |
| 76 | Low | 0.05 | High |
| 116 | High | 0.04 | High |
| 117 | Low | 0.1 | Low |
| 119 | High | 0.09 | Low |
| 121 | Low | 0.05 | High |
| 122 | Low | 0.1 | Low |
| 125 | High | 0.03 | High |
| 126 | Low | 0.11 | Low |
| 127 | High | 0.04 | High |
| 128 | Low | 0.06 | Low |
| 129 | Low | 0.11 | Low |
| 130 | High | 0.03 | High |
| 131 | High | 0.04 | High |
| 132 | Low | 0.03 | High |
| 135 | High | 0.11 | Low |

**Table 3:** Confusion matrix of aptamarker predictions versus Aβ42/Aβ40 ratio for low and high brain amyloid deposition.

|  | Predicted High | Predicted Low |
| --- | --- | --- |
| High | 11 | 4 |
| Low | 6 | 15 |

This confusion matrix outlines the performance of Aptamarker predictions in the context of brain amyloid deposition, with the Aβ42/Aβ40 ratio serving as the reference standard. A ratio less than 0.05 is considered indicative of high amyloid accumulation.

Where Predicted High (high brain amyloid deposition) and Predicted Low (low brain amyloid deposition) refer to Aptamarker test predictions of the amyloid ratio. This results in a sensitivity of 73%, specificity of 71% and overall accuracy of 72%.

Table 4 provides a comparison between the Aptamarker prediction and the designation of AD by the clinicians based on the full CSF analysis and a confusion matrix for summarising AD classifications are provided in Table 5. This table compares the predictive outcomes of Aptamarker testing with the clinical diagnosis of AD as determined by comprehensive cerebrospinal fluid (CSF) analysis.

**Table 4:** Comparison of aptamarker predictions with clinical diagnoses of AD based on CSF analysis.

| **Samples** | **Prediction** | **Diagnosis** |
| --- | --- | --- |
| 1 | Low | Depression |
| 4 | High | AD |
| 7 | Low | Normal |
| 8 | Low | AD and Vascular Dementia |
| 10 | Low | Depression |
| 11 | High | Depression |
| 14 | Low | Anti NMDA encephalitis |
| 15 | Low | Dementia by possible LATE TDP-43 |
| 16 | Low | Depression |
| 18 | Low | Frontotemporal dementia |
| 55 | High | Depression |
| 24 | High | AD (cortical posterior atrophy variant) |
| 29 | High | AD |
| 32 | High | AD (logopenic variant) |
| 37 | Low | Normal Pressure Hydrocephalus |
| 51 | High | AD |
| 42 | High | AD |
| 44 | Low | AD and Vascular Dementia |
| 45 | Low | Parkinson´s disease dementia |
| 56 | High | AD |
| 65 | High | AD |
| 76 | Low | AD |
| 116 | High | AD |
| 117 | Low | Depression |
| 119 | High | Depression |
| 121 | Low | AD |
| 122 | Low | Depression |
| 125 | High | AD |
| 126 | Low | Vascular Dementia |
| 127 | High | AD |
| 128 | Low | AD |
| 129 | Low | Depression |
| 130 | High | AD |
| 131 | High | AD |
| 132 | Low | AD |
| 135 | High | Lewy Body Dementia |

**Table 5:** Confusion matrix summarizing AD classifications based on Aptamarker predictions.

|  |  | Predicted brain amyloid |  |
| --- | --- | --- | --- |
|  |  | High | Low |
| Diagnosis | AD | 13 | 6 |
|  | Non AD | 4 | 13 |

The confusion matrix summarises the performance of Aptamarker predictions in the classification of AD. The sensitivity for this comparison was 68%, specificity 82% and overall accuracy was once again 75%. An overview of the diagnoses in relations to the Aptamarker predictions is provided in Figure 1.

**Figure 1:**
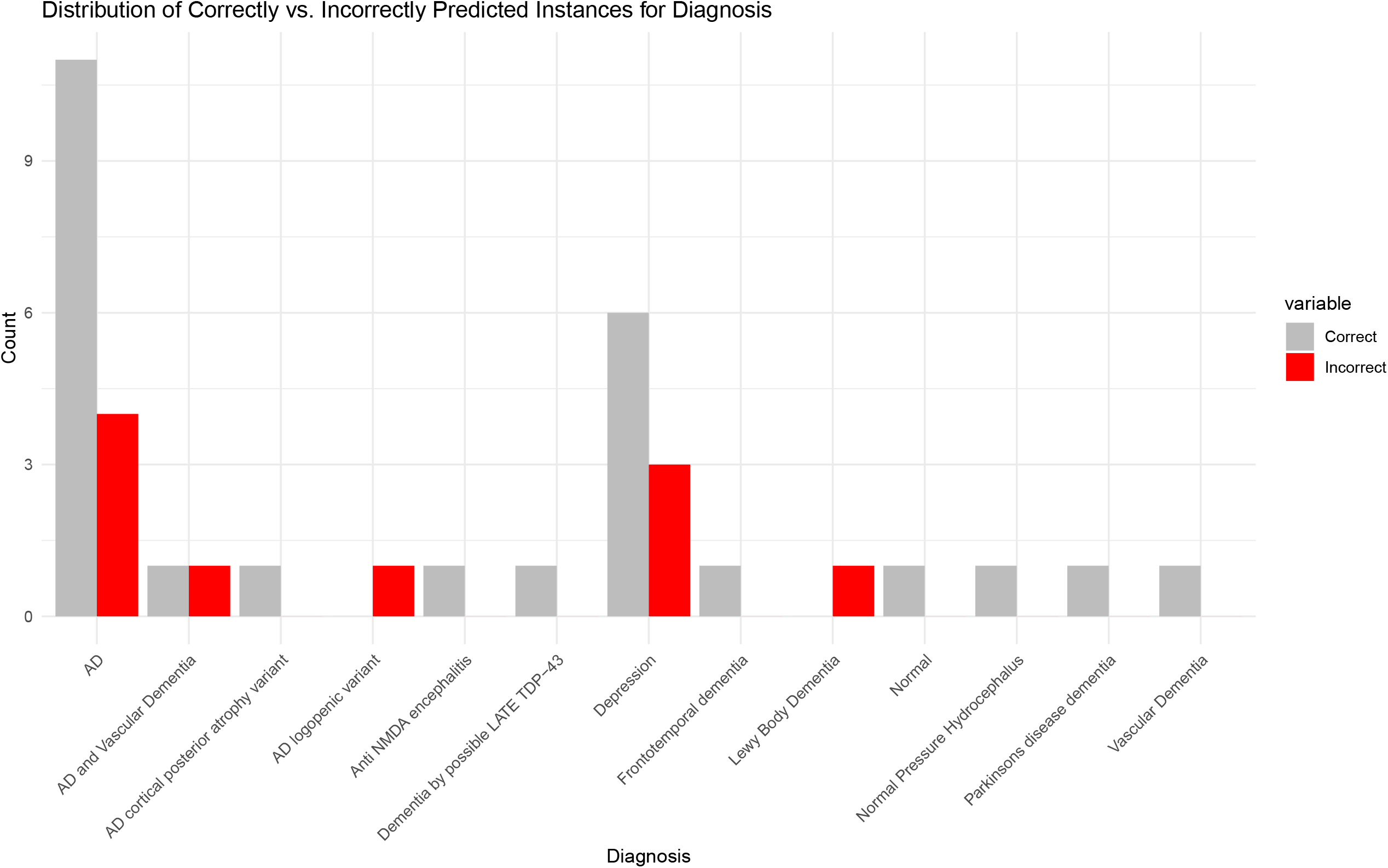
Distribution of mis-predicted individuals on the basis of clinical diagnosis. It is interesting to note that the three individuals predicted by the Aptamarker test as having high brain amyloid which did not have high brain amyloid all exhibited depression. This is too small a data set to speculate on any such correlations other than to note this observation for future studies.

## Discussion

The development of our predictive model was based on the analysis of 390 individuals from the AIBL cohort. It would be reasonable to expect some range of variation in predictive performance when analyzing a sample set that is less than 10% of the size of the original study. In this case however the observed results closely mimic those of the previous study, thus demonstrating the validity of this test.

From our results we draw the following conclusions.

1.) The gold standard for the confusion matrix in our algorithm development was PET scan analysis of the brains of the individuals involved, in this study the gold standard was CSF analysis. This represents a maintenance of predictive capacity with a change in gold standards meaning that our test is not limited to one gold standard versus the other. This demonstrates that our test is predicting brain amyloid deposition, independently of how this deposition is measured.

2.) The CHEDV study involved fresh blood extracted from patients in an ongoing clinical setting rather than samples that had been frozen for ten years or more (AIBL cohort). This means that the Aptamarkers are detecting molecules in the blood samples that are not significantly affected by storage conditions, or variations in the method of extraction. The CHEDV samples were subjected to at least three freeze/thaw cycles prior to analysis.

3.) The CHEDV study was constituted of individuals who exclusively identified themselves as Portuguese regarding ethnicity. This does not represent a broadening of ethnic diversity from the Australian study in terms of all individuals being Caucasian, and this is a gap that we intend to address in future studies. This does however represent a difference in lifestyle between Portugal and Australia of the individuals involved.

## Conclusion

We intend to pursue the application of this test as a first line test prior to CSF or alternative method of confirmative analysis. If an individual tests positive for brain amyloid deposition with the blood test, then this result would be confirmed by more sophisticated confirmatory tests. If individuals test negative, they will continue to be tested on an ongoing basis, but they would be examined by MRI to determine of vascular dementia and/or other causes.

## Data Availability

All data produced in the present study are available upon reasonable request to the authors

## List of abbreviations

AD: Alzheimer’s disease
AIBL: Australian imaging, biomarkers, and lifestyle
CHEDV: Centro Hospitalar de Entre Douro e Vouga
CSF: Cerebrospinal fluid
CT: Computed Axial Tomography
GNPs: Gold nanoparticles
IDT: Integrated DNA Technologies
MCI: Mild cognitive impairment
MRI: Magnetic resonance imagery
NFL: Neurofilament light chain
PBS: Phosphate buffer saline
PCR: Polymerase chain reaction
qPCR: Quantitative polymerase chain reaction
RTp: Relative template proportion
TCEP: Tris (2carboxyethyl) phosphine

## Declarations

### Ethics Approval and Consent to Participate

The research processes received ethical approval from the Ethics and Health Committee of Centro Hospitalar Entre o Douro e Vouga, Santa Maria da Feira, under the approval number n.53/2022. Informed consent was obtained from all participants or their legal representatives, who were informed about the study’s objectives, procedures, and potential risks in accordance with the Declaration of Helsinki. Eligible patients who underwent CSF analysis in the second semester of 2023 and fulfilled the inclusion criteria were consecutively invited to participate in the study.

### Revised Ethics of Data

All data utilized in this study have been anonymized at the source. We did not directly collect this information. Nonetheless, the personal data accessed cannot be used to identify specific individuals. Furthermore, all methods were performed in accordance with relevant guidelines and regulations, ensuring the integrity and confidentiality of the participants’ data.

## Availability of Data

The data underlying the figures and tables presented in this study are available at the following DOI: 10.6084/m9.figshare.24408451.

## Consent for publication

Not applicable.

## Author’s contributions

Gregory Penner: Conceived and designed the experiments, coordinated the collaboration between the two institutions, and was responsible for the final editing and approval of the manuscript.

Soizic Lecocq: Led the qPCR analysis, aptamer design, and antisense design. Assisted in writing and revising the manuscript.

Filipa Bastos, Mariana Silva, Rita Castro, João Cunha, Ana Cristina Silva, and Luis Ruano: Contributed to the patient recruitment, clinical diagnosis, and data collection at CHEDV. Collaborated in study design and defining inclusion criteria. Assisted in writing the clinical setting section.

Cathal Meehan: Responsible for data preprocessing and analysis from CHEDV at NeoNeuro and drafted and finalised the manuscript.

Luis Ruano: Also contributed to the study from an academic perspective due to affiliation with Universidade do Porto, providing insights into study design and data interpretation.

All authors were involved in reviewing the manuscript and approved the final version.

## Acknowledgements

The participation of all patients in this trial and their caregivers is greatly appreciated. The publication of the results will be shared with each of them. This project was financially supported by Roche Diagnostica, Portugal through their participation in the Building Tomorrow Together Initiative, coordinated by Beta -I, Portugal.

